# In-Hospital Mortality Trends Across the Pre-Pandemic, Pandemic, and Post-Pandemic Eras in Cardiovascular and Cerebrovascular Conditions: A Retrospective Cohort Study

**DOI:** 10.1101/2025.07.07.25331028

**Authors:** Swatam Jain, Pinak Shah, Alfred Danielian, Boone Singtong, Chris Aboujoude, Zade Zahlan, Rakahn Haddadin, Kush Kapadia

## Abstract

**Background:** The COVID-19 pandemic strained healthcare delivery, but its lasting impact on acute and chronic cardiovascular and cerebrovascular mortality remains unclear. We compared in-hospital mortality for ST-elevation myocardial infarction (STEMI), non-ST-elevation myocardial infarction (NSTEMI), ischemic stroke, and congestive heart failure (CHF) across pre-pandemic, pandemic, and post-pandemic eras.

**Methods:** We performed a retrospective cohort study of 38,735 U.S. hospital admissions (Jan 1, 2019–May 31, 2024) classified as pre-pandemic (Jan 2019–Mar 2020), pandemic (Mar 2020– May 2023), or post-pandemic (May 2023–May 2024). Cases were identified using ICD-10 codes. Multivariable logistic regression—adjusted for age, sex, race, diabetes, hypertension, chronic kidney disease, end-stage renal disease, COPD, and acute COVID-19 infection— assessed mortality odds across eras for each condition.

**Results:** STEMI (n=6,798) and stroke (n=18,142) mortality did not differ significantly across eras. NSTEMI (n=11,684) and CHF (n=8,775) mortality peaked during the pandemic (NSTEMI OR 1.47; CHF OR 1.53 vs post-pandemic; p<0.05) and declined markedly afterward. Chronic kidney disease (OR 1.41–1.75), end-stage renal disease (OR 2.28–3.40), and acute COVID-19 infection (OR 2.18–2.71) were independent predictors of higher mortality.

**Conclusions:** STEMI and stroke mortality remained stable across all three eras—likely due to established “code STEMI” and “code stroke” protocols. NSTEMI and CHF mortality peaked during the pandemic and improved post-pandemic, potentially reflecting better resourcing, enhanced care delivery, and widespread implementation of guideline-directed medical therapy. Sustained chronic disease management and emergency protocols are essential to optimize outcomes during and after healthcare crises.

## Introduction

The coronavirus disease 2019 (COVID-19) pandemic, caused by the novel severe acute respiratory syndrome coronavirus-2 (SARS-CoV-2), was first identified in Wuhan, China, in December 2019 and declared a global pandemic by the World Health Organization on March 11, 2020. (1) Over the next three years, hospitals were compelled to absorb unprecedented surges of critically ill COVID-19 patients while maintaining routine services. This dual burden strained bed capacity, depleted critical supplies, and exposed gaps in pandemic preparedness, even as personal protective equipment requirements and infection-control measures slowed emergency□department workflows. (2)

Cardiovascular emergencies proved especially vulnerable to these systemwide stresses. The management of acute coronary syndrome was likely impacted at various levels during the COVID-19 pandemic. Contributing factors included patients being more hesitant to call emergency medical services, reduced ambulance availability, longer emergency department wait times, and delays in performing percutaneous coronary intervention (PCI) due to the need for personal protective measures (3). Moreover, several studies have reported significantly increased in-hospital mortality for NSTEMI, acute decompensated heart failure, and ischemic stroke during the pandemic compared with pre-pandemic periods (4-6).

During the COVID-19 pandemic, the Society for Cardiovascular Angiography and Interventions (SCAI) reported that many patients perceived hospital visits as high-risk for SARS-CoV-2 transmission and feared infection more than a myocardial infarction. In the United Kingdom, late presentations of STEMI rose from 0% before the pandemic to 26% during it, leading to poorer clinical outcomes, increased mortality, and a higher incidence of cardiogenic shock (7,8). On 5 May 2023, the WHO declared that COVID-19 no longer constituted a Public Health Emergency of International Concern (9). Cardiovascular services immediately reset for a post-pandemic environment by replenishing critical supplies, refining triage algorithms, and bolstering staffing levels. Yet, data on cardiovascular outcomes in the post-pandemic period remain sparse in the literature. With patients’ renewed confidence in seeking urgent care and continued infection-control vigilance, acute cardiac presentations have returned to timelier levels, restoring full capacity for emergency interventions and improving clinical outcomes in the post-pandemic era.

We hypothesize that in-hospital mortality for patients with STEMI, NSTEMI, and heart failure will follow a “pandemic-peak” pattern, such that mortality is highest during the COVID-19 era, intermediate before the pandemic, and lowest after. During the pandemic, overwhelmed hospitals faced bed and staffing shortages and patients often delayed or avoided care, all of which likely drove the highest mortality rates. In contrast, the pre-pandemic period—while not burdened by COVID-related resource constraints—represents a baseline of standard care with intermediate outcomes. We further anticipate that the post-pandemic era will demonstrate reduced mortality, reflecting improved allocation of resources, better-equipped facilities, and a rebound in provider and system responsiveness—manifested as greater awareness, prompt triage, and more timely interventions.

## Materials and Methods

### Data Source

Data were obtained from the HCA Healthcare enterprise database in a multicenter retrospective analysis of admissions across all HCA-affiliated hospitals in the United States. The dataset contained fully de-identified patient information and did not require direct patient interaction or additional institutional review board approval. All de-identification processes adhered to applicable patient privacy regulations.

### Study Population

A total of 38,735 patients aged 18 to 90 years were included. Eligible patients had an admission diagnosis of STEMI, NSTEMI, ischemic stroke, or CHF, as defined by ICD-10 codes (see Supplementary Table 1). Patients under 18 or over 90 years of age were excluded.

### Study Design

This retrospective chart review analyzed admissions between January 1, 2019, and May 31, 2024. Four separate multivariable logistic regression models were constructed—one for each condition (STEMI, NSTEMI, CHF, and ischemic stroke)—to assess the odds of in-hospital mortality across three defined periods: pre-pandemic, pandemic, and post-pandemic. Additional models assessed individual predictors of mortality, including demographic and clinical variables such as age, sex, race, diabetes, hypertension, chronic kidney disease (CKD), end-stage renal disease (ESRD), chronic obstructive pulmonary disease (COPD), and acute COVID-19 infection.

### Study Cohorts

Patients were grouped based on the date of admission into three timeframes: Pre-pandemic (January 1, 2019–March 10, 2020), Pandemic (March 11, 2020–May 4, 2023), and Post-pandemic (May 5, 2023–May 31, 2024), in accordance with World Health Organization declarations [1,9]. Patient cohorts were balanced based on key characteristics, including age, sex, race, and comorbidities.

### Study Outcome and Variables

The primary outcome was in-hospital mortality, defined as death during the index hospitalization (binary: yes/no). Independent variables included demographics (age, sex, race) and comorbid conditions: hypertension, diabetes mellitus, CKD, ESRD, COPD, and COVID-19 infection. All variables were coded dichotomously (present or absent) and were incorporated into propensity score models. A full list of ICD-10 codes is provided in Supplementary Table 2.

### Statistical Analysis

To reduce confounding and ensure comparability between eras, 1:1 propensity score matching was performed using key covariates: age, sex, race, hypertension, diabetes, CKD, ESRD, COPD, and COVID-19 infection. Matched cohorts were used to assess differences in in-hospital mortality across the three pandemic-defined eras.

Post-matching, four separate logistic regression models were developed—one per condition (STEMI, NSTEMI, stroke, and CHF)—to calculate adjusted odds ratios (ORs) with 95% confidence intervals (CIs) for in-hospital mortality. Results were interpreted in the context of clinical relevance and statistical significance.

## Results

### ST□Elevation Myocardial Infarction (STEMI)

Among 6,798 STEMI admissions, 558 patients (8.2%) died in-hospital. In multivariable regression, neither the pandemic nor pre-pandemic era was significantly associated with mortality relative to the post-pandemic period (p > 0.50). Female sex (OR 1.211; 95% CI 1.003–1.460; p=0.0467), hypertension (OR 1.573; 95% CI 1.245–1.987; p=0.0001), diabetes mellitus (OR 1.616; 95% CI 1.338–1.952; p<0.0001), end-stage renal disease (OR 2.645; 95% CI 1.874–3.733; p<0.0001), and COVID-19 infection (OR 2.378; 95% CI 1.512–3.740; p=0.0002) were all significantly associated with higher mortality. (See table 2 and 3)

**Table 1.**
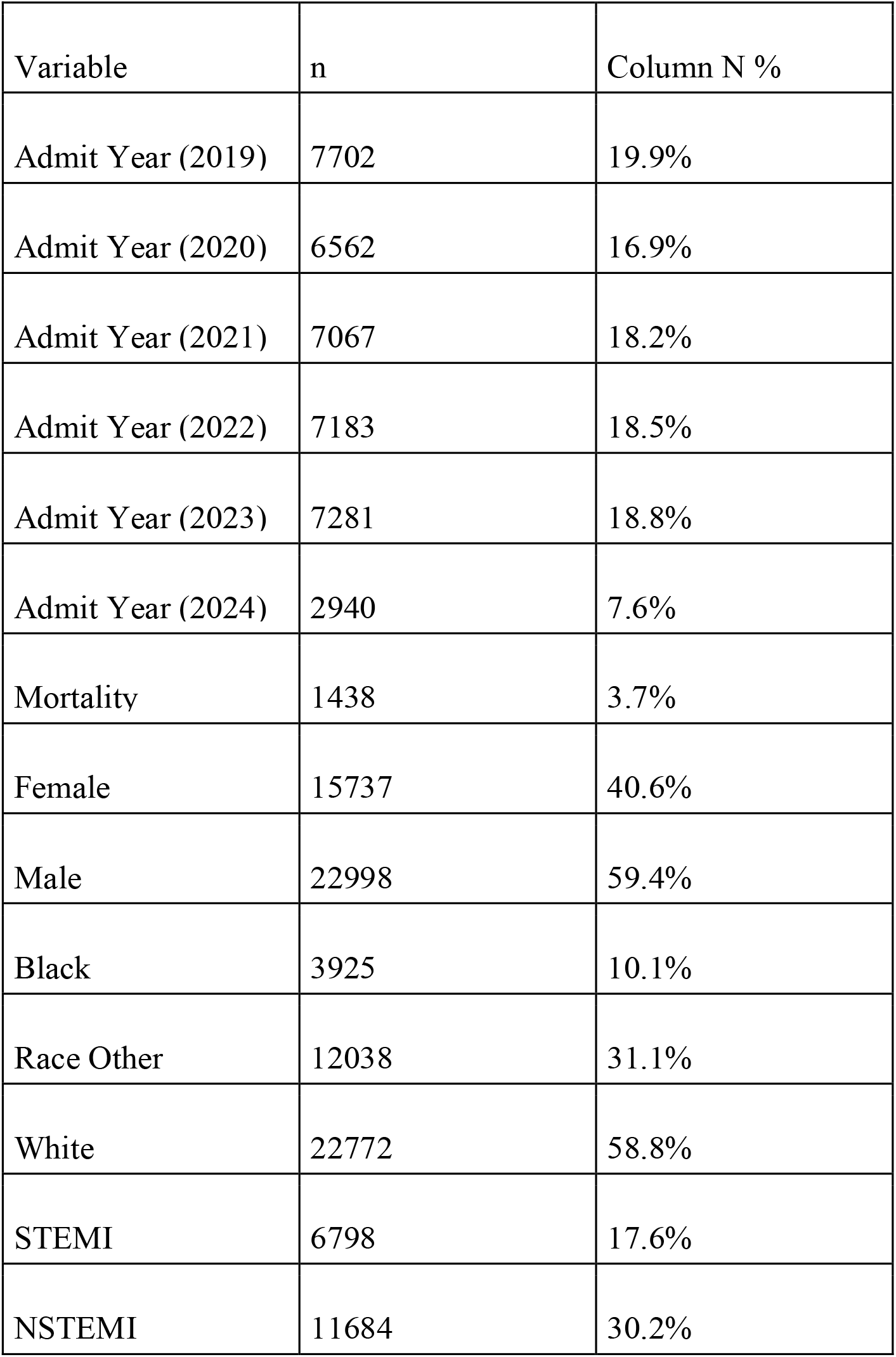
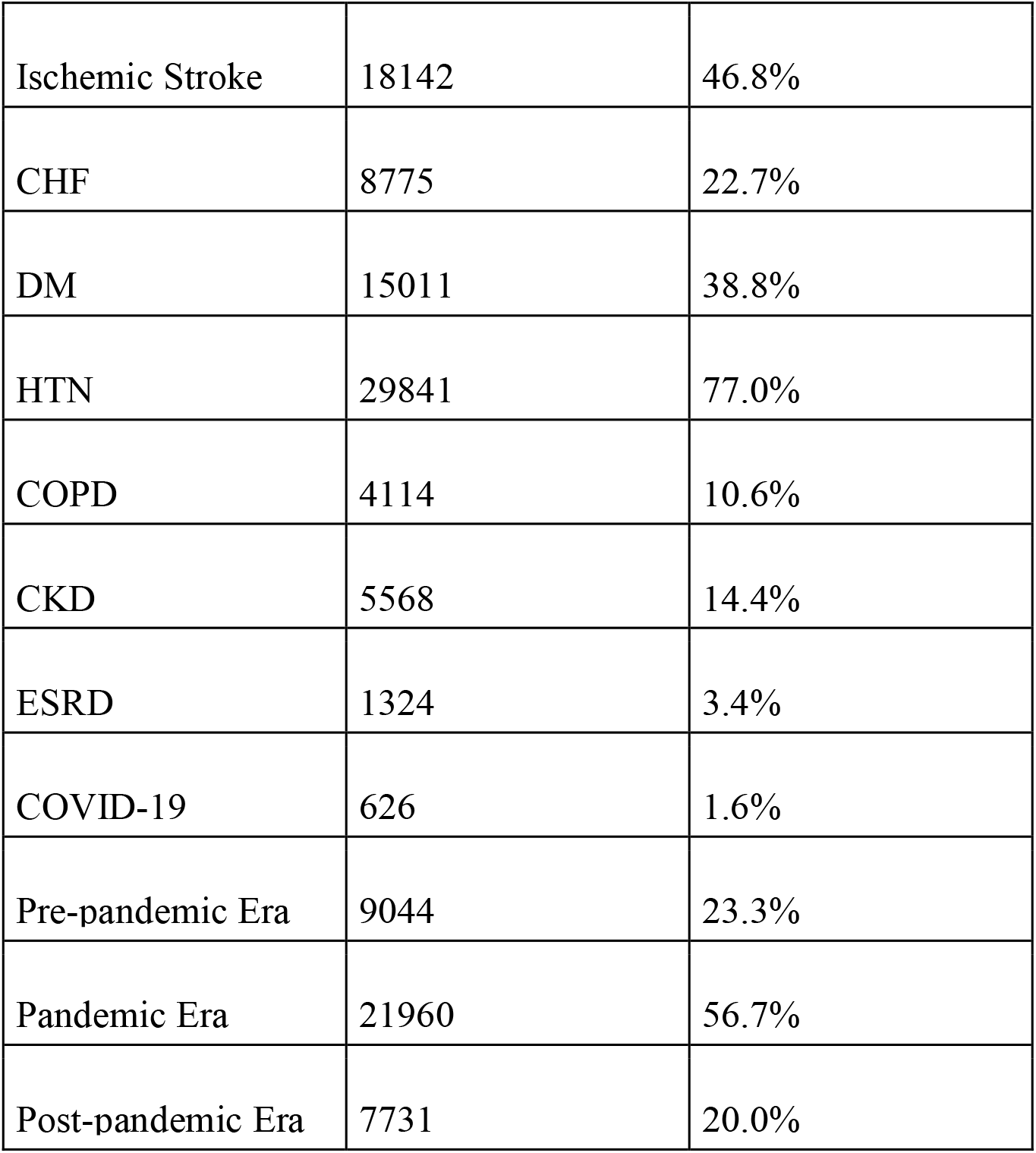
Descriptive Statistics for the Overall Cohort (n = 38,735)

| Variable | n | Column N % |
| --- | --- | --- |
| Admit Year (2019) | 7702 | 19.9% |
| Admit Year (2020) | 6562 | 16.9% |
| Admit Year (2021) | 7067 | 18.2% |
| Admit Year (2022) | 7183 | 18.5% |
| Admit Year (2023) | 7281 | 18.8% |
| Admit Year (2024) | 2940 | 7.6% |
| Mortality | 1438 | 3.7% |
| Female | 15737 | 40.6% |
| Male | 22998 | 59.4% |
| Black | 3925 | 10.1% |
| Race Other | 12038 | 31.1% |
| White | 22772 | 58.8% |
| STEMI | 6798 | 17.6% |
| NSTEMI | 11684 | 30.2% |
| Ischemic Stroke | 18142 | 46.8% |
| CHF | 8775 | 22.7% |
| DM | 15011 | 38.8% |
| HTN | 29841 | 77.0% |
| COPD | 4114 | 10.6% |
| CKD | 5568 | 14.4% |
| ESRD | 1324 | 3.4% |
| COVID-19 | 626 | 1.6% |
| Pre-pandemic Era | 9044 | 23.3% |
| Pandemic Era | 21960 | 56.7% |
| Post-pandemic Era | 7731 | 20.0% |

**Table 2:** Adjusted Odds Ratios for Era Comparisons Across Cohorts.

| Era Comparison | STEMI<br>OR (95% CI; p-value) | NSTEMI<br>OR (95% CI; p-value) | Ischemic stroke<br>OR (95% CI; p-value) | CHF<br>OR (95% CI; p-value) |
| --- | --- | --- | --- | --- |
| Pre- vs Post-Pandemic | 0.97 (0.74–1.28; p = 0.84) | 1.47 (1.07–2.02; p = 0.017) | 1.273 (0.983–1.649; p = 0.0670) | 1.53 (1.13–2.08; p = 0.0058) |
| Pandemic vs Post-Pandemic | 1.05 (0.83–1.33; p = 0.71) | 1.33 (1.00–1.76; p = 0.051) | 1.209 (0.966 – 1.513; p= 0.0981) | 1.33 (1.01–1.74; p = 0.040) |
| Pre- vs Pandemic | 0.93 (0.64–1.34; p ≈ 0.68) | 1.11 (0.73–1.69; p ≈ 0.63) | 0.949 (0.778 – 1.158; p = 0.6086) | 1.16 (0.77–1.74; p ≈ 0.48) |

**Table 3:**
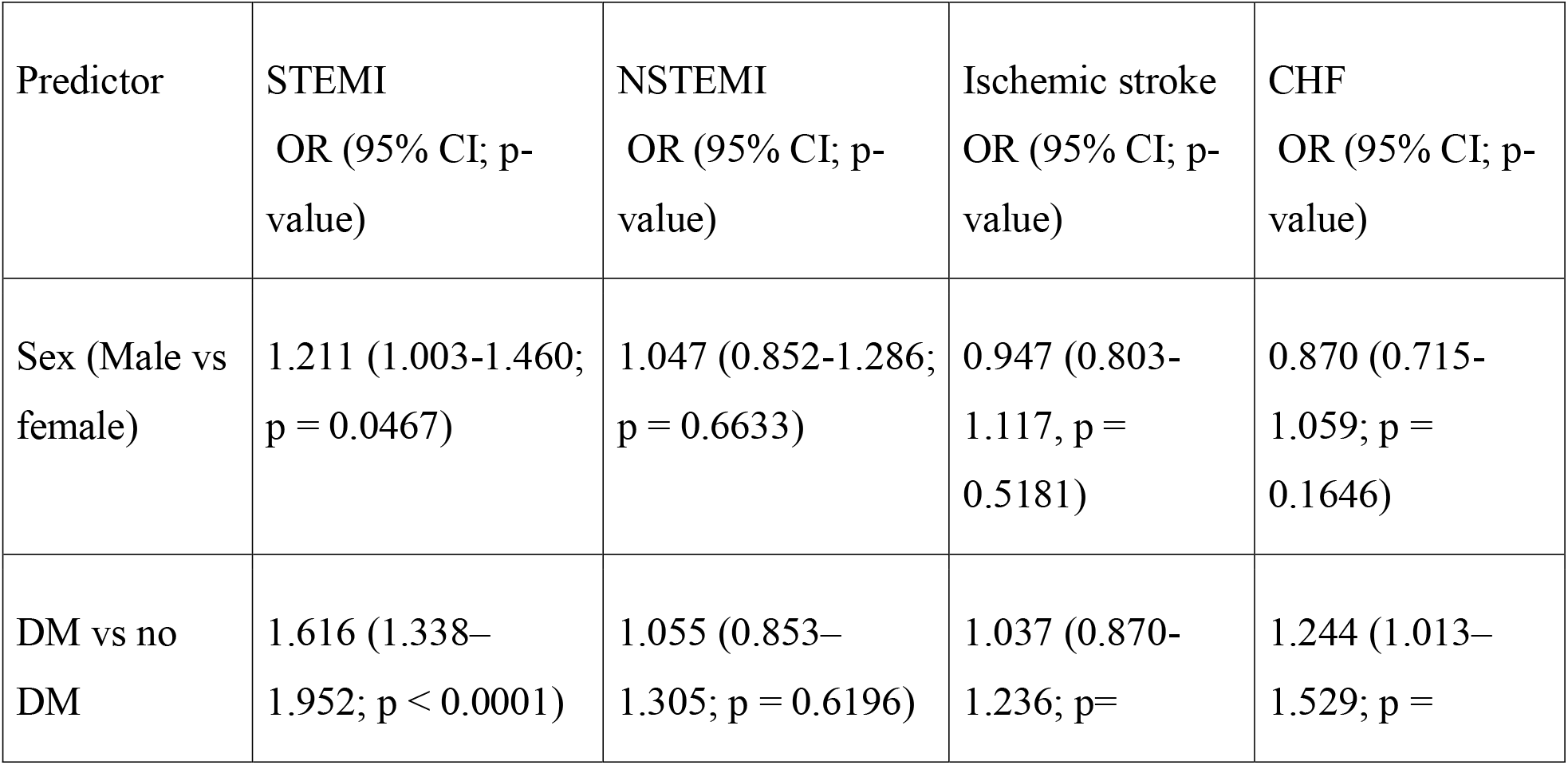
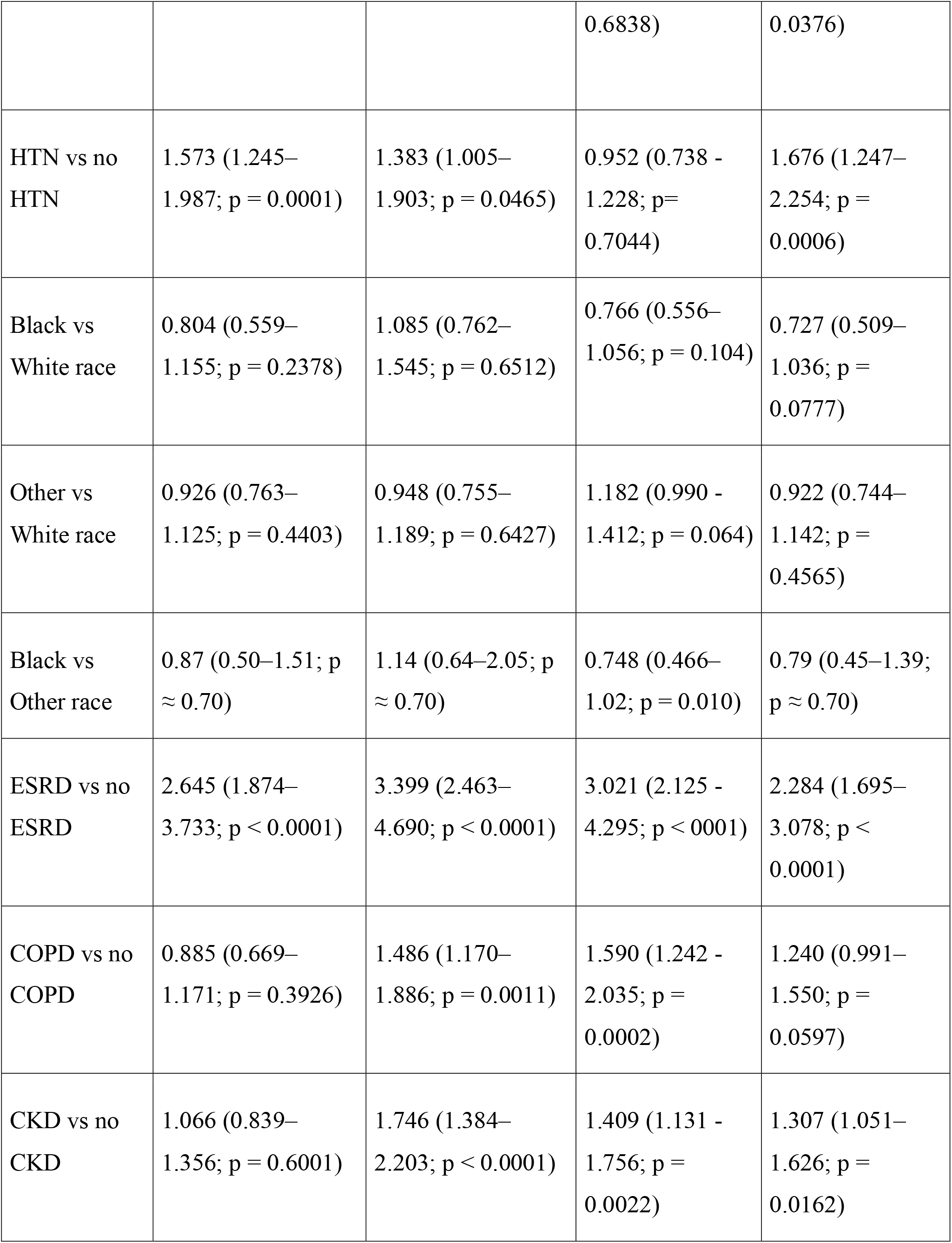
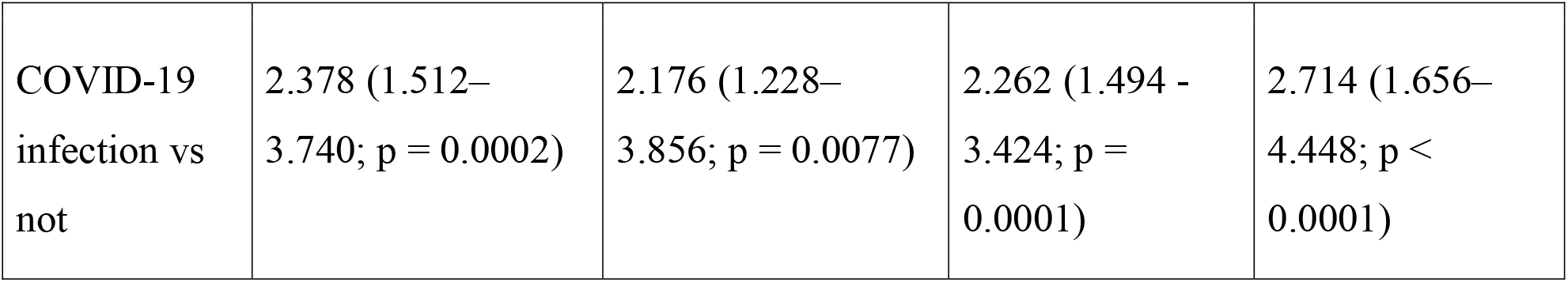
Adjusted Odds Ratios for Key Covariates in the Analyses.

### Non–ST□Elevation Myocardial Infarction (NSTEMI)

Of 11,684 NSTEMI admissions, 416 (3.6%) resulted in in-hospital death. Compared to the post-pandemic era, pre-pandemic admissions had a 47% higher mortality risk (OR 1.468; 95% CI 1.070–2.015; p=0.0173), and pandemic-era admissions showed a borderline increase (OR 1.325; 95% CI 0.999–1.759; p=0.0511). Hypertension (OR 1.383; 95% CI 1.005–1.903; p=0.0465), chronic kidney disease (OR 1.746; 95% CI 1.384–2.203; p<0.0001), end-stage renal disease (OR 3.399; 95% CI 2.463– 4.690; p<0.0001), COPD (OR 1.486; 95% CI 1.170–1.886; p=0.0011), and COVID-19 infection (OR 2.176; 95% CI 1.228–3.856; p=0.0077) were significant mortality predictors. (See table 2 and 3).

### Ischemic Stroke

Among 18,142 ischemic stroke admissions, 607 patients (3.3%) died in-hospital. Mortality did not significantly differ between pandemic (OR 1.209; 95% CI 0.966– 1.513; p=0.0981), pre-pandemic (OR 1.273; 95% CI 0.983–1.649; p=0.0670), and post-pandemic periods. Chronic kidney disease (OR 1.409; 95% CI 1.131–1.756; p=0.0022), end-stage renal disease (OR 3.021; 95% CI 2.125–4.295; p<0.0001), COPD (OR 1.590; 95% CI 1.242–2.035; p=0.0002), and COVID-19 infection (OR 2.262; 95% CI 1.494–3.424; p=0.0001) were all associated with significantly higher mortality. (See table 2 and 3).

### Congestive Heart Failure (CHF)

In the CHF cohort of 8,775 admissions, 465 patients (5.3%) died. Compared with the post-pandemic era, both the pandemic (OR 1.325; 95% CI 1.012–1.735; p=0.0404) and pre-pandemic periods (OR 1.534; 95% CI 1.132–2.079; p=0.0058) were associated with higher in-hospital mortality. Hypertension (OR 1.676; 95% CI 1.247–2.254; p=0.0006), diabetes mellitus (OR 1.244; 95% CI 1.013–1.529; p=0.0376), chronic kidney disease (OR 1.307; 95% CI 1.051–1.626; p=0.0162), end-stage renal disease (OR 2.284; 95% CI 1.695–3.078; p<0.0001), and COVID-19 infection (OR 2.714; 95% CI 1.656–4.448; p<0.0001) were all significant predictors. COPD showed a trend toward higher mortality (OR 1.240; 95% CI 0.991–1.550; p=0.0597). (See table 2 and 3).

### Summary

Across all conditions, STEMI and stroke mortality remained unchanged across eras, while NSTEMI and CHF mortality peaked during the pandemic and improved post-pandemic. Chronic kidney disease, end-stage renal disease, and COVID-19 infection consistently predicted higher in-hospital mortality.

## Discussion

Our study evaluated how the COVID-19 pandemic influenced in-hospital mortality for STEMI, NSTEMI, ischemic stroke, and CHF patients across the United States. Contrary to the initial hypothesis, mortality did not uniformly increase during the pandemic era. Instead, STEMI and ischemic stroke mortality remained remarkably stable across the three defined periods, whereas NSTEMI and CHF mortality peaked during the pandemic, declined thereafter, and was lowest in the post-pandemic era.

These condition-specific differences highlight the nuanced effects of the pandemic on cardiovascular and cerebrovascular outcomes.(10) For STEMI and ischemic stroke, the preservation of acute-care protocols—such as code STEMI and code stroke pathways—likely enabled continued access to timely and effective interventions, buffering these conditions from systemic healthcare disruptions.(11) In contrast, NSTEMI and CHF care—which relies more heavily on coordinated outpatient follow-up and timely medical optimization—was significantly impacted by deferred care, reduced access to providers, and hesitancy to seek medical attention during the pandemic.

Our results are consistent with previous literature showing that STEMI mortality remained stable during the pandemic due to protected emergency systems and revascularization pathways. For example, a study from Japan reported no decline in PCI rates for ACS nor increase in mortality during the pandemic, and our findings align with these results. (12) However, our data also show that STEMI patients with concurrent COVID-19 infection had more than double the odds of in-hospital death, reinforcing the independent risk posed by SARS-CoV-2. (13,14)

The significant rise in NSTEMI and CHF mortality during the pandemic likely reflects reduced utilization of guideline-directed medical therapy (GDMT), delays in seeking care, and impaired access to cardiovascular diagnostics. Similarly, studies have shown worse mortality in the pandemic compared to pre-pandemic times. (4-6, 15) Key medications—such as sacubitril/valsartan and SGLT2 inhibitors—were more widely adopted in the post-pandemic era, which may explain the mortality improvements observed in our data. (16-19) Moreover, studies have shown that limited use of invasive strategies in NSTEMI patients and reduced outpatient support contributed to worse outcomes during the pandemic. (20, 21)

During the pandemic, resource limitations, overwhelmed healthcare systems, and reduced outpatient follow-up impaired the longitudinal management of chronic cardiovascular conditions. In the post-pandemic period, replenished supplies, streamlined protocols, and strengthened staffing enhanced provider responsiveness and system robustness—bolstering outcomes in both NSTEMI and CHF cohorts. Simultaneously, patients became more health-conscious, presenting earlier in their disease course and adhering more closely to guideline-directed medical therapies—particularly for chronic issues like heart failure—further driving the observed improvements in CHF outcomes.

In contrast, ischemic stroke outcomes remained stable across all three time periods. Several international and U.S. studies support this finding, citing uninterrupted stroke code protocols and the prioritization of acute neurovascular interventions such as thrombolysis and mechanical thrombectomy. This suggests that hospitals-maintained stroke treatment infrastructure even amid significant systemic pressures. (22-24)

Independent predictors of mortality across all cohorts included end-stage renal disease, chronic kidney disease, and COVID-19 infection. These findings are consistent with existing literature and emphasize the additive burden of chronic illness and acute infection in driving worse outcomes. (25-28) Interestingly, race was not a significant predictor of mortality in our adjusted analyses, suggesting equitable in-hospital care delivery, though this does not account for disparities in access prior to admission. (29)

## Strengths and Limitations

This study leverages a large, nationally representative cohort with standardized data collection across multiple facilities and time periods. Adjustment for key comorbidities strengthens the validity of our findings. Notably, this is among the first studies to analyze post-pandemic mortality outcomes for both cardiovascular and cerebrovascular diseases.

However, limitations include the retrospective design and reliance on administrative data, which may introduce misclassification or residual confounding. We did not distinguish between elective and emergent admissions, nor could we account for outpatient treatment quality or socioeconomic determinants of care. Finally, COVID-19 testing rates and treatment protocols may have varied over time and between sites.

## Conclusion

In this large retrospective cohort analysis of patients hospitalized with STEMI, NSTEMI, ischemic stroke, or CHF, in-hospital mortality for STEMI and ischemic stroke remained stable across the pre-, pandemic, and post-pandemic eras. In contrast, NSTEMI and CHF mortality peaked during the pandemic and subsequently declined. These findings reflect the resilience of acute care systems and the critical role of timely interventions and GDMT in optimizing patient outcomes. Chronic kidney disease, end-stage renal disease, and acute COVID-19 infection were the most consistent predictors of in-hospital mortality across conditions.

## Data Availability

All data produced in the present study are available upon reasonable request to the authors

## Clinical Perspectives

- **Preserved Acute Care Protocols Mitigated Mortality in STEMI and Stroke:** Despite the systemic disruptions of the pandemic, in-hospital mortality for STEMI and ischemic stroke remained stable across all eras, likely due to protected code STEMI/stroke pathways and uninterrupted access to emergent interventions.
- **NSTEMI and CHF Outcomes Were Vulnerable to Pandemic Disruptions:** In-hospital mortality for NSTEMI and CHF peaked during the pandemic, highlighting the sensitivity of these conditions to delays in care, reduced outpatient follow-up, and impaired access to guideline-directed therapy.
- **Post-Pandemic Improvements Reflect System Recovery and Patient Behavior Shifts:** The decline in NSTEMI and CHF mortality in the post-pandemic era likely reflects improved healthcare delivery, broader adoption of evidence-based therapies, and increased patient engagement in timely care-seeking and medication adherence.

## Acknowledgments

The authors declare no acknowledgments.

## Supplementary Material

**Supplementary table 1:** ICD -10 codes used for Inclusion criteria

| Admission Diagnosis | ICD-10 Code(s) |
| --- | --- |
| STEMI | I21 (excluding I21.4) |
| NSTEMI | I21.4 |
| Ischemic Stroke | I63 |
| Congestive Heart Failure | I50 |

**Supplementary table 2:** ICD -10 codes used for Categorical Variables

| Condition | ICD-10 Code(s) |
| --- | --- |
| Diabetes mellitus | E08–E13 |
| Ischemic stroke | I63 |
| Hypertension | I10–I16 |
| Chronic obstructive pulmonary disease (COPD) | J41, J42, J43, J44 |
| Chronic kidney disease (CKD) (excluding ESRD) | N18 (exclude N18.6) |
| End-stage renal disease (ESRD) | N18.6 |
| COVID-19 | U07.1 |

